# Flat Foot and Lower Back Pain: An Association and Implications for Treatment

**DOI:** 10.1101/2025.05.15.25327574

**Authors:** Manogna Sagiraju, Raghav Prasad

## Abstract

**Background:** Lower back pain (LBP) is a leading cause of disability worldwide, with multifactorial etiology making it challenging to treat. Flatfoot is an often overlooked condition that might significantly contribute to the LBP burden.

**Objective:** The primary objective is to establish flat foot as a risk factor for LBP. The study aims to determine the prevalence of flatfoot in patients with LBP compared to those without.

**Methods:** This study was conducted at a private clinic in Chicago who were randomly selected and screened for flatfoot and LBP. The prevalence of flatfoot among patients with LBP was measured, and the odds ratio was calculated.

**Results:** Out of the patients with LBP, 27.3% had flatfoot, compared to only 9.5% among those without LBP. The study found that patients with LBP were 3.54 times more likely to have flatfoot. This association was particularly strong in non-obese, male patients over sixty years of age.

**Conclusion:** The study suggests that flatfoot is a risk factor for LBP. This finding calls for a change in the approach to LBP assessment, including early screening and treatment of asymptomatic flatfoot to prevent complications later in life.

## Introduction

Flat foot or pes planus is a syndrome combining multiple static and dynamic deformities with partial or complete collapse of the medial longitudinal arch (MLA) [1] [2], which causes the entire sole to contact the ground in a near-complete or complete manner. Additionally, it is characterized by the abduction of the forefoot, internal rotation, plantar flexion of the talus, and calcaneal eversion [3].

The prevalence of adult flat foot is 15–25%, based on a study in 2020 [4], while another study in Massachusetts revealed a prevalence of 19% [5] and 26.6% in another study in Spain [6]. In comparison, lower back pain (LBP) has a worldwide prevalence of 18% [7]. In 1998, LBP was responsible for $90.7 billion in healthcare expenditures in the USA [8]. This number has only increased since then, as LBP remained the leading cause of YLDs globally from 1990 to 2017 [9] and is expected to increase to 843 million patients by 2025 [10].

Understanding the significance of LBP, Lancet recently advocated for governments, policymakers, and the wider community to take measures in addressing the burden of lower back pain [10]. High-income countries have developed better healthcare systems to manage this increasing burden. For low-and middle-income countries, healthcare infrastructure will be unable to cope and will face a more significant challenge in managing the growing LBP burden in the coming years.

A systematic review in 2013 found that despite limited research on the connection between flat foot and lower back pain, early studies showed at least a partial causative relationship between the two [11]. In the military, applicants with flat foot have historically been rejected because of the risk of developing foot and back pain [12]. It can increase the risk of hallux valgus, patellofemoral pain syndrome, and LBP [13].

The prevalence of LBP among participants with flat foot was 65.9%, compared with the 32.8% prevalence of LBP among those without flat foot, as seen in a study in Saudi Arabia [14]. One of the drawbacks of that study was that all the flat foot data was self-reported which can lead to recall bias. In comparison this study depends on clinical records or clinician evaluation to diagnose flat foot providing a more accurate estimate of the prevalence of flat foot.

There has been a substantial growth in prescription of opioid medication for chronic musculoskeletal pain including LBP [15] which has led to increased cases of substance use disorder and opioid related overdose [16]. Establishment of flat foot as an independent factor for LBP can help us move away from a nonspecific and blanket approach of prescribing pain medications. Instead it could promote implementing lifestyle modifications and specific measures that target the root cause of LBP. Also, proactively managing adults with flat foot prior to LBP symptoms can lead to a reduction in the burden of the disease.

This study aimed to determine the prevalence of flat foot in patients with and without lower back pain and to ascertain an association between the two in Chicago, Illinois, USA.

## Methods

The study was conducted at a clinic in Chicago, Illinois, in the month of April 2023. Ethical Clearance was obtained, and written informed consent was obtained from all participants. Patients were randomly selected from those who arrived at the clinic for regular health check-up.

Patients with recent trauma, previous back surgery, malformation of the foot, substance use disorder, osteoporosis, spinal stenosis, fracture of the spine, neoplasia, infection of the spine (osteomyelitis, epidural abscess), inflammatory arthritis, prostatitis, endometriosis, nephrolithiasis, pyelonephritis, perinephric abscess, aortic aneurysm, pancreatitis, or penetrating ulcer were excluded from the study.

General demographic data of patient like age and sex was obtained from patients via self-reporting. Patients were classified based on age into two groups, sixty years and below and above sixty years. Patient was asked about smoking history and was considered a smoker if they had smoked in the last 15 years. Clinical records were checked for history of comorbidities such as hypertension, diabetes and dyslipidemia. Patient weight and height were measured and body mass index (BMI) was calculated. Patients were classified as obese if BMI was greater than 30 and non-obese if BMI was less than 30.

Patient was then asked about history of LBP and if present then the duration of symptoms. Case files were reviewed to check for previous diagnosis of flat foot. If no such diagnosis existed then, using the visual method flat foot is evaluated. Two independent clinicians had to agree to make the diagnosis of flat foot.

All statistical analysis was performed on JASP (version 0.18) and R (version 4.3.2), p-values of less than 0.05 were considered statistically significant. The baseline characteristics and the prevalence of flat foot were compared between the LBP and non LBP group. The Shapiro-Wilk Test for normality was performed for all continuous variables. The normally distributed data was compared using students *t* test and non-normally distributed data utilised the Mann-Whitney U test. For categorical variables chi-squared test were used. The effect of flat foot on lower back pain was investigated using logistic regression analysis.

## Results

A total of 215 patients were studied: 110 with LBP and 105 without LBP. The age, sex and BMI of patients with and without LBP were not significantly different. There was no significant difference in the presence of comorbidities (hypertension, dyslipidemia and diabetes) and smokers in both the groups. The average duration of symptoms in patients with LBP was 6.4 years. The details are listed in **Table 1**.

**Table 1.**
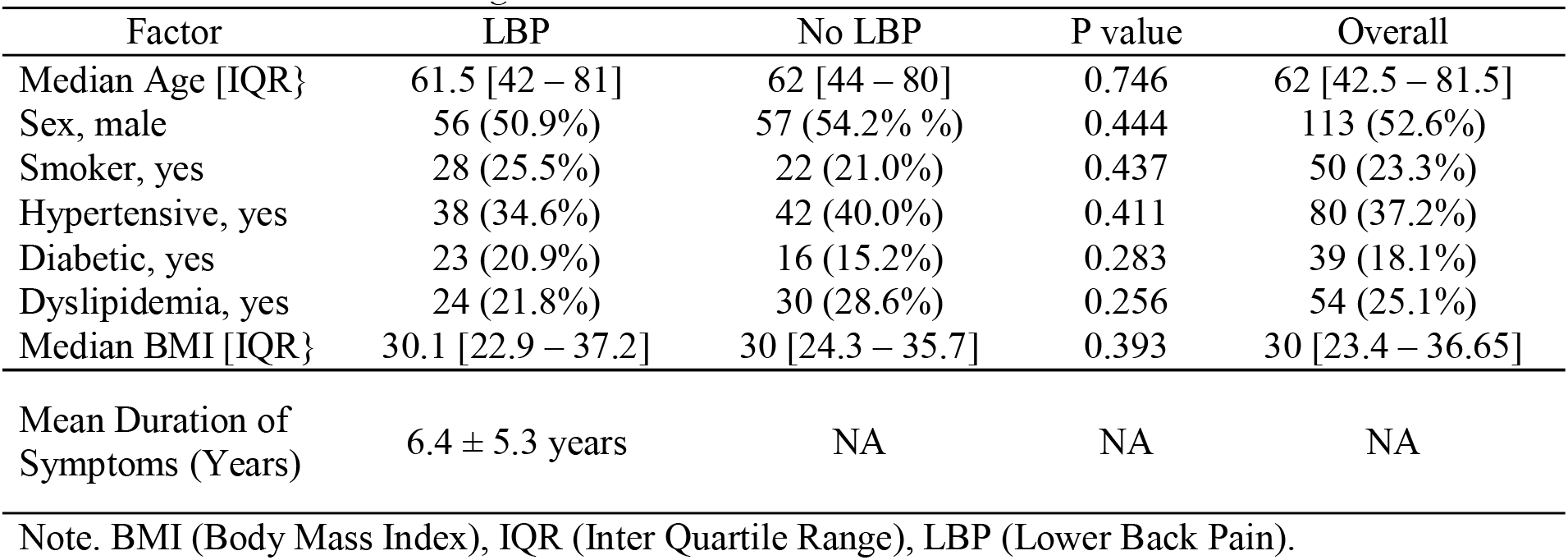
Patient distribution Based on Age and Sex.

Of the 215 patients, 40 had flat foot, thus accounting for a prevalence of 18.6%. When divided based on presence and absence of LBP the prevalence was 27.3% and 9.5% respectively. Patients were further divided based on sex, age and obesity in **Table 2**. The higher prevalence of flat foot was found in patients with lower back pain especially among males (35.8%), patients aged at or less than sixty years (34.0%) and non-obese patients (34.0%). Thus, the highest prevalence of flat foot is expected in a male, non-obese patient at or under the age of sixty with lower back pain which came up to be 62.5%.

**Table 2.**
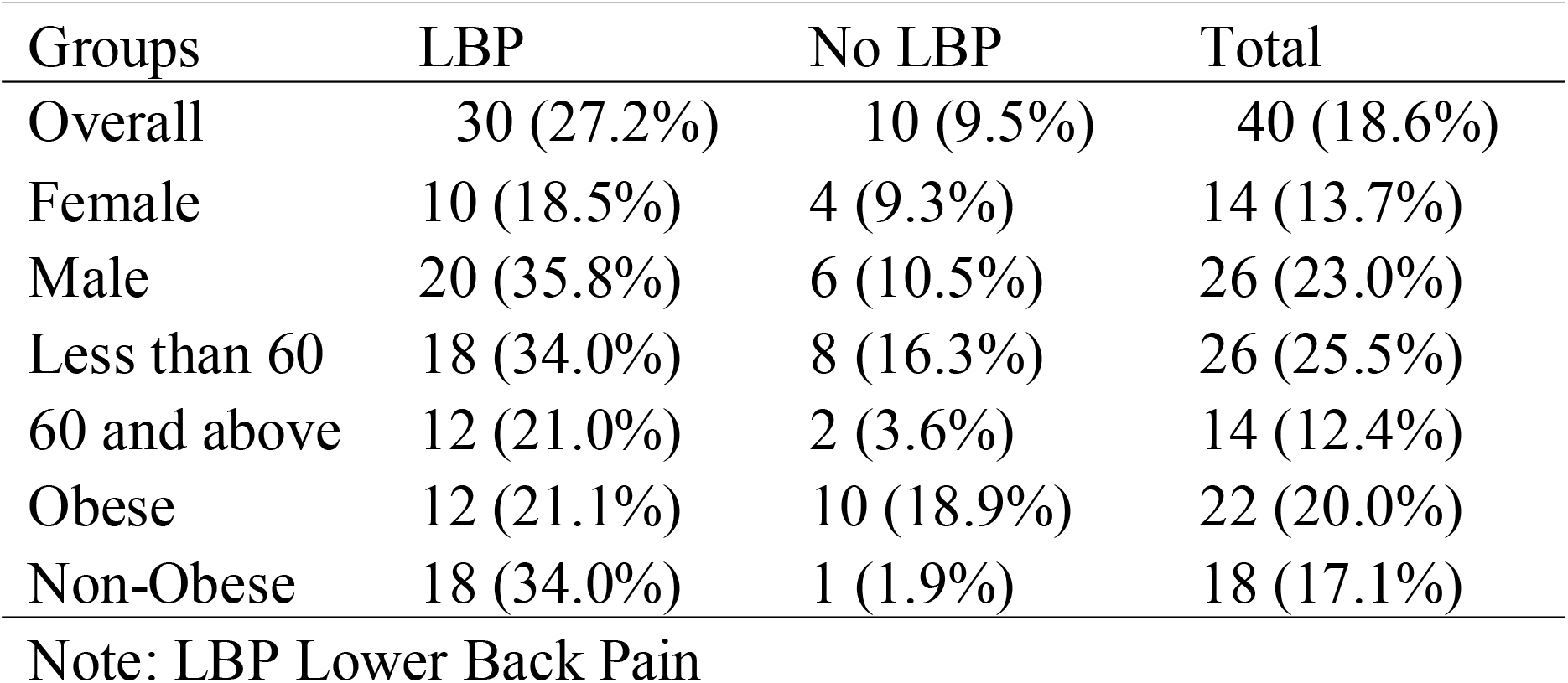
Prevalence of flat foot.

The Odds ratio in all patients with LBP for flat foot came to 3.542 which was statistically significant. When classified based on sex, no significant association was seen in female patients while in male patients an odds ratio of 4.658 (CI: 1.602 - 15.635). On classifying based on age no significant association was seen in patients aged at or less than that sixty years (p > 0.05). Whereas in patients aged greater than sixty years of age an odds ratio of 7.09 was seen (CI: 1.464 – 68.320). When divided based on BMI, no significant association was seen in obese patients (p > 0.05) while in non-obese patients an odds ratio of 25.59 (CI: 3.709 – 1107.382) with p <0.01 is established. Details are mentioned in **Table 3**.

**Table 3.**
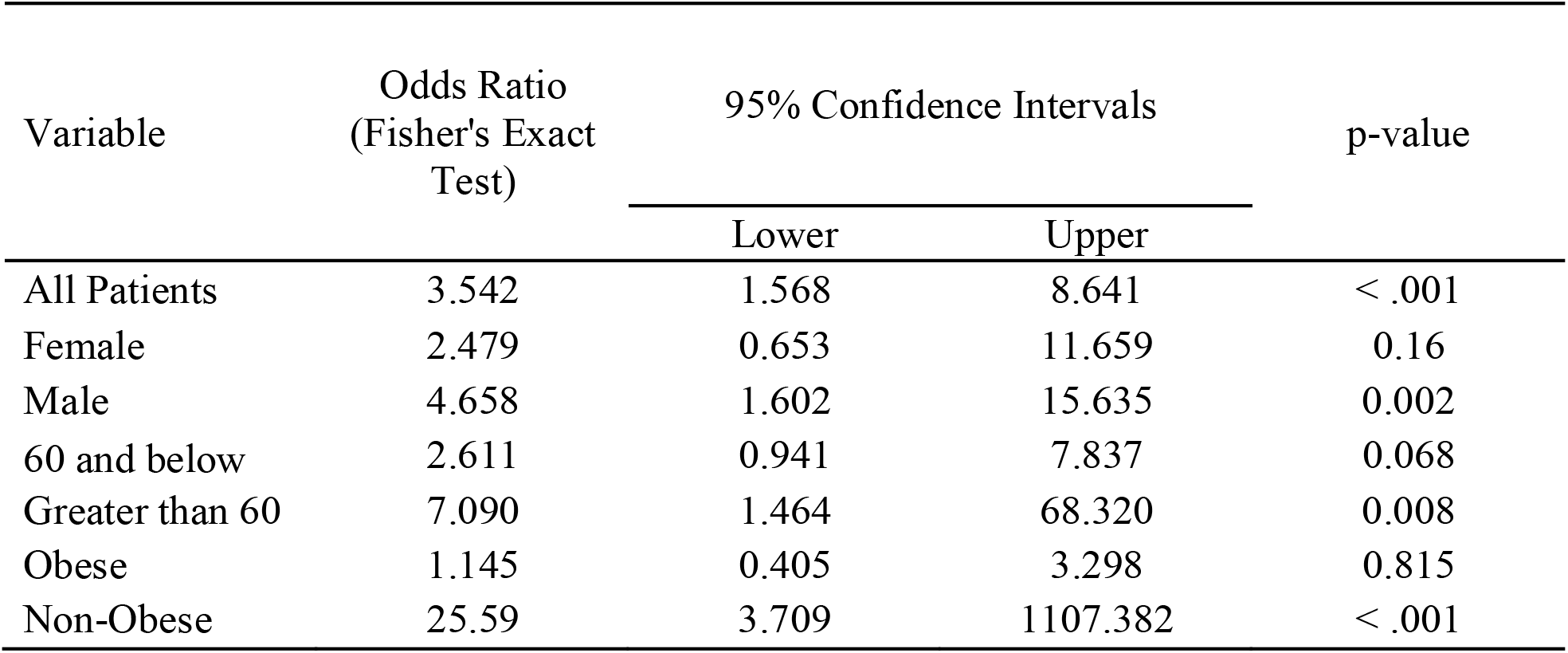
Odds Ratio Between LBP and Flat Foot.

Thus, the patients most likely to have flat foot as a risk factor for lower back pain are male, non-obese patients above the age of sixty years. This group consists of only 28 patients making the sample size insufficient.

## Discussion

The medial longitudinal arch is the differentiating factor of the foot in humans compared with primates and is responsible for humans walking upright. The arches of the foot are strongest on the plantar surface. The structure and dynamicity of the arch are essential for shock absorption, body weight transmission, and as a lever to propel the body forward during locomotion [12].

Flat foot in children are normal and expected until age six due to increased ligament laxity, which declines with age [17]. One of the factors responsible for pediatric flat foot is the use of footwear at an early age, as observed in a study conducted among school-going children in India [18]. When the condition persists or develops after skeletal maturity, it is known as Adult Flat Foot [19]. The effect has not been studied on the incidence of flat foot in adults and was not examined in this study either. Future studies should look into it to promote footwear that provides young feet with the necessary support and structure for optimal growth and mobility. We can promote healthy foot development in children by emphasizing arch development in the design.

Flat foot can be further classified as flexible and rigid feet. The flexible phenotype is one in which the arch deforms only on weight-bearing and restores itself on cessation of the same. The rigid type has collapsed arches irrespective of the weight-bearing status of the foot [20]. In this study, we did not differentiate between the two types of flat foot. This is necessary as treatment of asymptomatic flexible flat foot is usually deferred as compared to prompt management of rigid flat foot. Further studies are needed to assess the effects of flexible and rigid flat foot on LBP as it can influence the current management guidelines.

In our study the prevalence of flat foot in our population of 18.6% is within the range of 15 to 26.6% as seen in previous studies [4-7]. The study established a significant association between flat foot and LBP. An odds ratio of 3.542 means that patients with LBP were 3.5 times more likely to have flat foot. The next step is to perform prospective longitudinal studies to establish the degree of risk of LBP in patients with flat foot. Currently, small clinical trials have already established a reduction in LBP after prescribing foot orthoses compared to placebo in patients with flat foot [21].

A possible explanation for the association between flat foot and LBP is the compensatory change in posture for better dynamic balance, which can alter the pelvic alignment [22] and the electromyographic activity of the erector spinae and gluteal muscles during walking [23]. These maladaptations can have a damaging effect on the lower extremities and spine [24].

Another theory is that of the “anatomy train.” It states that any tension in a particular part of the anatomy train will result in detrimental effects on the other parts [25]. The foot is part of the anatomy train, involving the entire lower extremity and ending in the lower back; thus, any malformation in the foot leads to tension in the lower back [11,26]. This tension can eventually lead to degenerative changes in the spine, as observed in a study in 2021, in which the incidence rates of degenerative changes in the spine in the flat and non-flat foot cohorts were 16 and 11 per 10,000-person months, respectively [27].

In this study, the average duration of LBP symptoms was 6.2 years. This meant that most patients undergo extensive testing to establish the etiology of the back pain, leading to increased healthcare expenditure. In addition, the study found that a significant association existed only in patients greater than sixty years of age which could point towards an insidious process of disease progression. So, a possible method to decrease the burden and healthcare costs is by early screening of adults and management of flat foot before it causes symptomatic back pain. The large time period required to observe the effect of flat foot makes cohort studies and clinical trials less feasible, thus making case control studies such as this essential in improving our understanding of the disease.

In this study no, significant association was established in female patients while in male patients even though the association was significant the confidence intervals were very wide. It was expected that females would have a greater association as they are at a higher risk of LBP than males with flat foot due to increased static anterior pelvic tilt, internal hip rotation, and dorsal spine inclination [13,28]. A study in 2012 in Nigeria showed a higher prevalence of flat foot and LBP in females [29]. This difference could not be assessed in this study due to the lack of a significant sample size.

The prevalence of flat foot and LBP is higher in individuals with a higher body mass index [30]. This could explain the lack of a significant association of flat foot with LBP in obese patients as it acts as a confounding factor. Yet studies have shown that increased physical activity has aided in the improvement of foot structure and function among obese people [31], which can potentially lead to improvement in LBP symptoms.

The strength of the study lies in the fact that the diagnosis of flat foot was made by trained clinicians in comparison to previous studies where patients self-reported their flat foot status which improves accuracy. It also reduces the indigence of recall bias when reporting about their LBP symptoms. Even though not recorded the study was conducted at a clinic in Chicago with a diverse patient population allowing generalization of the results on a worldwide basis.

This study utilized the visual method for the diagnosis of flatfoot. The visual method is less reliable than the Navicular Drop method [12,32] but is less time-consuming and more convenient for patients. In addition, we did not measure the height of the Medial Longitudinal Arch, which can help determine the degree of flatfoot [33] and has been shown to influence the magnitude of acceleration at the lumbar spine during running [34].

Therefore, even though this study helps establish an association, further studies are needed to understand better the role of flat foot in the incidence of LBP. A better understanding can help promote changes, such as changes in guidelines regarding when and which type of footwear children can wear. It can introduce universal screening for flat foot in adults. Although most cases of flat foot are asymptomatic when discovered, they can cause problems for the individual, leading to unnecessary testing and treatment. Therefore, the management guidelines can be changed.

## Conclusion

This study demonstrated flat foot as a risk factor for chronic lower back pain with an odds ratio of 3.542. With an estimated 843 million cases of LBP by 2025, there is a need for early screening and management of cases of flat foot to reduce the burden of LBP. Potential measures such as specific footwear and lifestyle modifications into the treatment plan to benefit the patient. This will also help curb over prescription and dependence on opioids for long-term management of LBP. Further studies are needed to determine the amount of risk associated with flat foot and clinical trials to study the benefits of management of flat foot in decreasing the incidence and treatment of cases of lower back pain.

## Data Availability

All data produced in the present study are available upon reasonable request to the authors

## Acknowledgement

The authors have no acknowledgments.

## Author Contributions Statement

Manogna Sagiraju – Conceived and designed the study, collected the data, reviewed the manuscript and approved the final version.

Dr Raghav Prasad-Performed the analysis, wrote and reviewed the manuscript and approved the final version.

